# Durable Humoral and Cellular Immune Responses Following Ad26.COV2.S Vaccination for COVID-19

**DOI:** 10.1101/2021.07.05.21259918

**Authors:** Dan H. Barouch, Kathryn E. Stephenson, Jerald Sadoff, Jingyou Yu, Aiquan Chang, Makda Gebre, Katherine McMahan, Jinyan Liu, Abishek Chandrashekar, Shivani Patel, Mathieu Le Gars, Anne Marit de Groot, Dirk Heerwegh, Frank Struyf, Macaya Douoguih, Johan van Hoof, Hanneke Schuitemaker

**Affiliations:** Center for Virology and Vaccine Research, Beth Israel Deaconess Medical Center, Boston, MA, USA; Janssen Vaccines & Prevention, Leiden, The Netherlands; Janssen Research & Development, Beerse, Belgium

## Abstract

Interim immunogenicity and efficacy data for the Ad26.COV2.S vaccine for COVID-19 have recently been reported^1-3^. We describe here the 8-month durability of humoral and cellular immune responses in 20 individuals who received one or two doses of 5×10^10^ vp or 10^11^ vp Ad26.COV2.S and in 5 participants who received placebo^2^. We evaluated antibody and T cell responses on day 239, which was 8 months after the single-shot vaccine regimen (N=10) or 6 months after the two-shot vaccine regimen (N=10), although the present study was not powered to compare these regimens^3^. We also report neutralizing antibody responses against the parental SARS-CoV-2 WA1/2020 strain as well as against the SARS-CoV-2 variants D614G, B.1.1.7 (alpha), B.1.617.1 (kappa), B.1.617.2 (delta), P.1 (gamma), B.1.429 (epsilon), and B.1.351 (beta).

---

Antibody responses were detected in all vaccinees on day 239 (Fig. 1A). Median WA1/2020 receptor binding domain (RBD)-specific binding antibody titers were 645, 1772, 1962, and 1306 on days 29, 57, 71, and 239, respectively. Median WA1/2020 pseudovirus neutralizing antibody titers were 272, 169, 340, and 192 on days 29, 57, 71, and 239, respectively (Fig. 1A, upper panels), and were comparable when restricted to individuals who received the single-shot vaccine regimen (Fig. S1). Three Ad26.COV2.S vaccine recipients showed a sharp increase in antibody responses during this time period; one individual developed breakthrough SARS-CoV-2 infection and two received mRNA vaccines. Excluding these three participants, antibody responses were relatively stable over 8 months with only a 1.8-fold reduction of median neutralizing antibody titers between peak responses on day 71 and the durability timepoint day 239.

**Figure 1.**
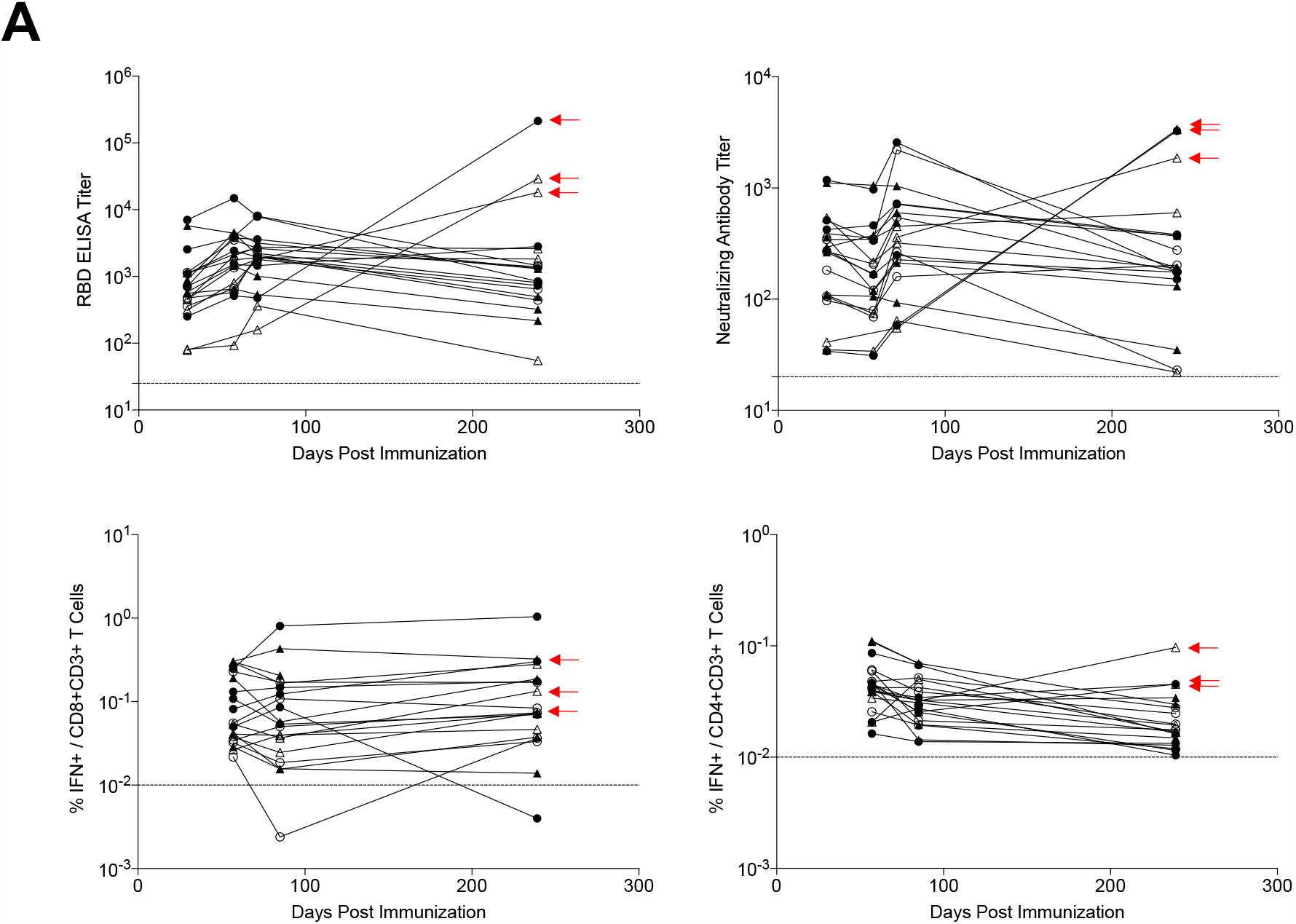
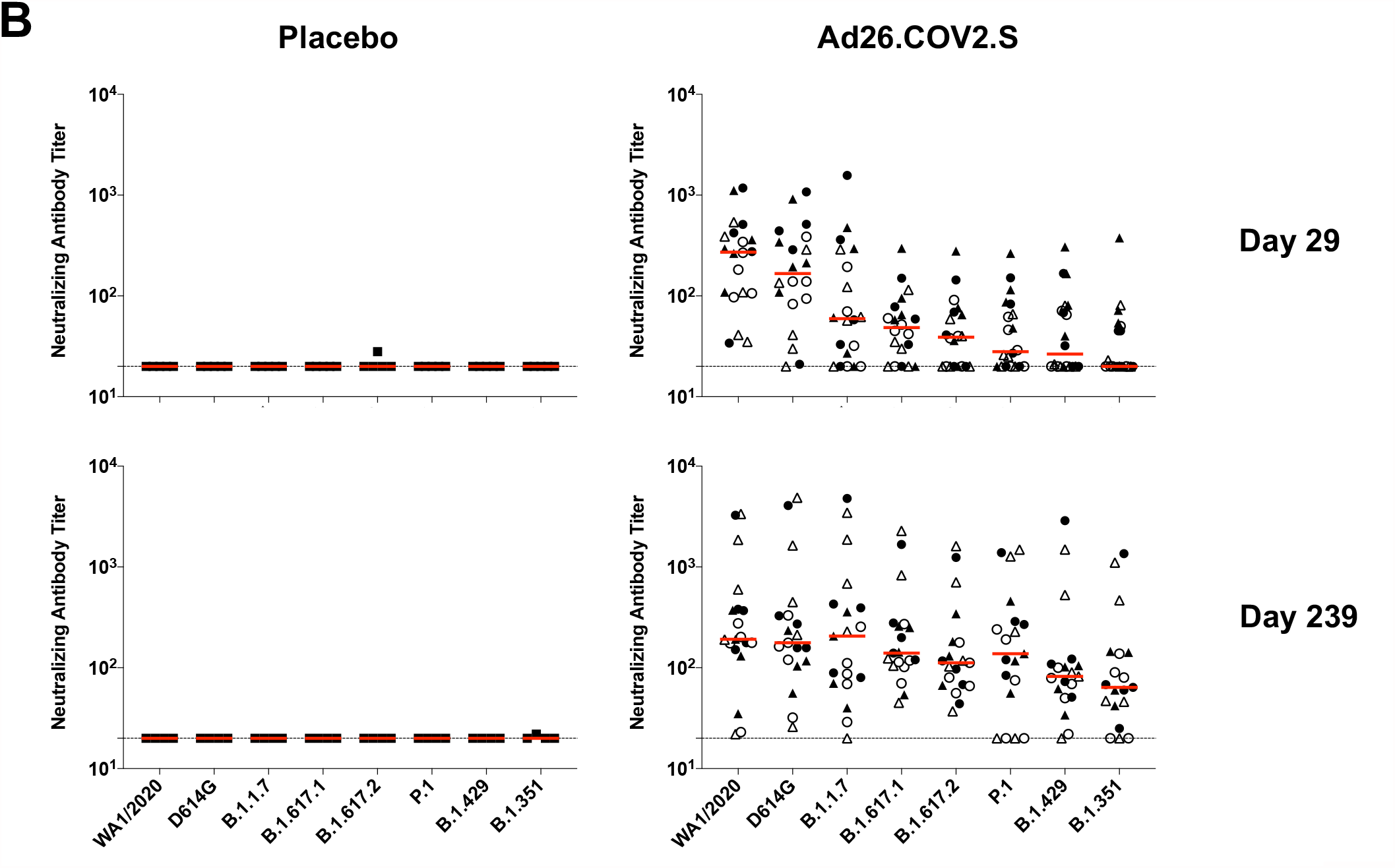
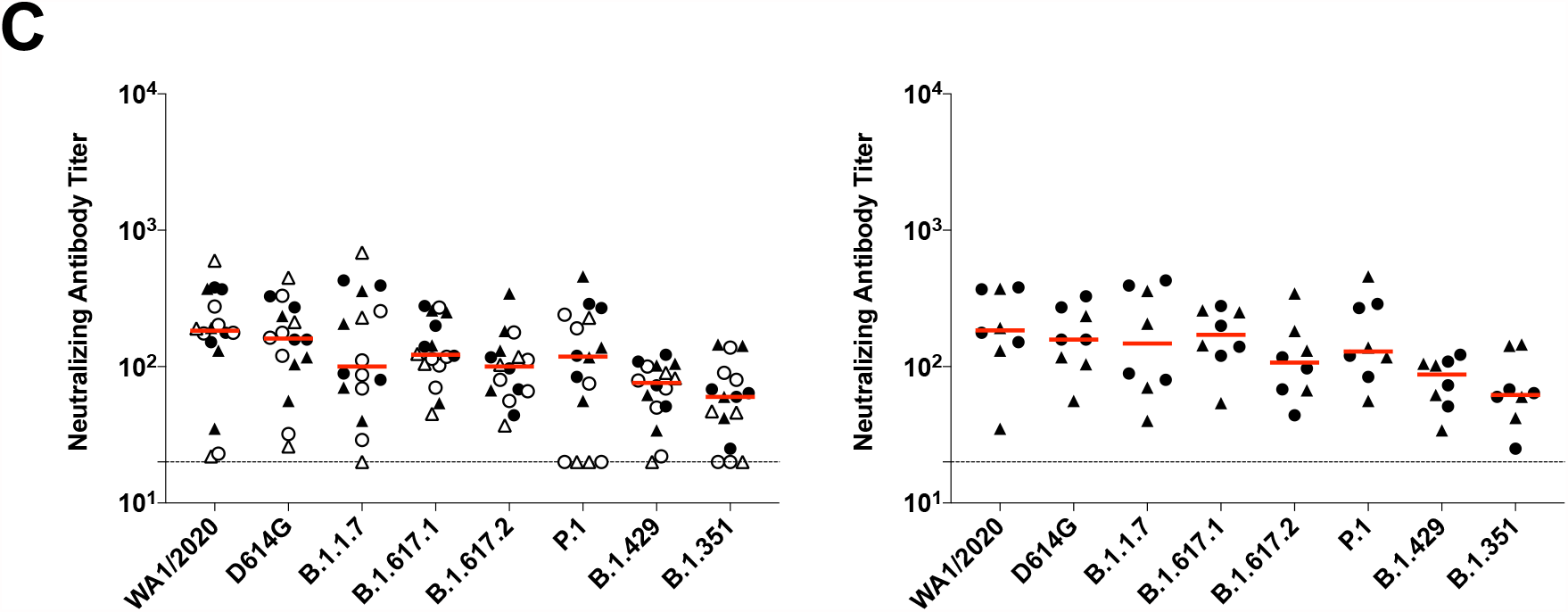
Durability of humoral and cellular immune responses following Ad26.COV2.S vaccination. (A) SARS-CoV-2 WA1/2020 receptor binding domain (RBD)-specific binding antibodies by ELISA, pseudovirus neutralizing antibody assays, and spike-specific CD8+ and CD4+ T cell responses by intracellular cytokine staining assays on days 29, 57, 71 or 85, and 239. Red arrows highlight three individuals who developed breakthrough SARS-CoV-2 infection (filled circle; N=1) or who received mRNA vaccines (open triangles; N=2) between days 71 and 239. (B) Pseudovirus neutralizing antibody assays against the parental WA1/2020 strain as well as the SARS-CoV-2 variants D614G, B.1.1.7 (alpha), B.1.617.1 (kappa), B.1.617.2 (delta), P.1 (gamma), B.1.429 (epsilon), and B.1.351 (beta) on days 29 and 239. (C) Left, pseudovirus neutralizing antibody assays on day 239 following Ad26.COV2.S vaccination excluding the three individuals who developed breakthrough SARS-CoV-2 infection or who received mRNA vaccines. Right, pseudovirus neutralizing antibody assays on day 239 also restricted to individuals who received single-shot Ad26.COV2.S vaccination. Red bars reflect median responses. Dotted lines reflect lower limits of quantitation based on the WA1/2020 assay. Filled squares, placebo; filled circles, 10^11^ vp (single dose); open circles, 10^11^ vp (two dose); filled triangles, 5×10^10^ vp (single dose); open triangles, 5×10^10^ vp (two dose). For the two-dose vaccine, immunizations were on Day 1 and Day 57.

On day 29, median neutralizing antibody titers showed a >13-fold reduction to the B.1.351 variant compared with WA1/2020 (Fig. 1B). On day 239, however, median neutralizing antibody titers showed a more modest 3-fold reduction to the B.1.351 variant compared with WA1/2020 (Fig. 1B). Excluding the three individuals who developed breakthrough SARS-CoV-2 infection or who received mRNA vaccines, and restricted to individuals who received the single-shot vaccine regimen, median neutralizing antibody titers on day 239 were 184, 158, 147, 171, 107, 129, 87, and 62 against the SARS-COV-2 variants WA1/2020, D614G, B.1.1.7 (alpha), B.1.617.1 (kappa), B.1.617.2 (delta), P.1 (gamma), B.1.429 (epsilon), and B.1.351 (beta), respectively (Fig. 1C). These data suggest an expansion of neutralizing antibody breadth with improved coverage of SARS-CoV-2 variants over time, including increased neutralizing antibody titers against these variants of concern.

Spike-specific IFN-*γ* CD8+ and CD4+ T cell responses were evaluated by intracellular cytokine staining assays and also showed durability and stability over this time period (Fig. 1A, lower panels). Median CD8+ T cell responses were 0.0545%, 0.0554%, and 0.0734% on days 57, 85, and 239, respectively. Median CD4+ T cell responses were 0.0435%, 0.0322%, and 0.0176% on days 57, 85, and 239, respectively.

These data show that the Ad26.COV2.S vaccine elicited durable humoral and cellular immune responses with minimal decline for at least 8 months following immunization. In addition, we observed an expansion of neutralizing antibody breadth against SARS-CoV-2 variants over this time period, including against the more transmissible B.1.617.2 (delta) variant and the partially neutralization resistant B.1.351 (beta) and P.1 (gamma) variants, suggesting maturation of B cell responses even without further boosting. The durability of immune responses elicited by Ad26.COV2.S is consistent with the durability reported for an Ad26-based Zika vaccine^4^. Longitudinal antibody responses to mRNA COVID-19 vaccines have also been reported for 6 months but with more rapid decline kinetics^5^. The durability of humoral and cellular immune responses following Ad26.COV2.S vaccination with increased neutralizing antibody responses to SARS-CoV-2 variants over time, including after single-shot vaccination, further support the use of the Ad26.COV2.S vaccine for the global COVID-19 pandemic.

## Data Availability

Requests for access to the study data can be submitted to D.H.B.

## ACKNOWLEDGMENTS

We acknowledge support from Janssen Vaccines & Prevention BV, the Ragon Institute of MGH, MIT, the Massachusetts Consortium on Pathogen Readiness (MassCPR), the Musk Foundation, the National Institutes of Health (CA260476). This project has been funded in part with federal funds from the Office of the Assistant Secretary for Preparedness and Response, Biomedical Advanced Research and Development Authority, under Other Transaction Agreement HHSO100201700018C.

## DATA SHARING STATEMENT

Requests for access to the study data can be submitted to D.H.B.

## SUPPLEMENTAL APPENDIX

### SUPPLEMENTARY METHODS

#### Pseudovirus-based neutralization assay

The SARS-CoV-2 pseudoviruses expressing a luciferase reporter gene were generated as we have previously described. Briefly, the packaging plasmid psPAX2 (AIDS Resource and Reagent Program), luciferase reporter plasmid pLenti-CMV Puro-Luc (Addgene), and Spike protein expressing pcDNA3.1-SARS CoV-2 SΔCT of variants were co-transfected into HEK293T cells by lipofectamine 2000 (ThermoFisher). Pseudoviruses of SARS-CoV-2 variants were generated by using the Spike from the WA1/2020 strain (Wuhan/WIV04/2019, GISAID accession ID: EPI_ISL_402124), D614G mutation, B.1.1.7 variant (GISAID accession ID: EPI_ISL_601443), B.1.617.1 variant (GenBank Accession ID: QTS25314.1), B.1.617.2 variant (GenBank Accession ID: QTW89558.1), P.1 variant (GISAID accession ID: EPI_ISL_792683), B.1.429 variant (GISAID accession ID: EPI_ISL_824730), or B.1.351 variant (GISAID accession ID: EPI_ISL_712096). The supernatants containing the pseudotype viruses were collected 48 h post-transfection, which were purified by centrifugation and filtration with 0.45 µm filter. To determine the neutralization activity of the plasma or serum samples from participants, HEK293T-hACE2 cells were seeded in 96-well tissue culture plates at a density of 1.75 × 10^4^ cells/well overnight. Three-fold serial dilutions of heat inactivated serum or plasma samples were prepared and mixed with 50 µL of pseudovirus. The mixture was incubated at 37°C for 1 h before adding to HEK293T-hACE2 cells. 48 h after infection, cells were lysed in Steady-Glo Luciferase Assay (Promega) according to the manufacturer’s instructions. SARS-CoV-2 neutralization titers were defined as the sample dilution at which a 50% reduction in relative light unit (RLU) was observed relative to the average of the virus control wells.

#### ELISA

WA1/2020, B.1.1.7, and B.1.351 RBD-specific binding antibodies were assessed by ELISA. Briefly, 96-well plates were coated with 2µg/ml RBD protein in 1X DPBS and incubated at 4°C overnight. After incubation, plates were washed once with wash buffer (0.05% Tween 20 in 1 X DPBS) and blocked with 350 µL Casein block/well for 2-3 h at room temperature. After incubation, block solution was discarded and plates were blotted dry. Serial dilutions of heat-inactivated serum diluted in casein block were added to wells and plates were incubated for 1 h at room temperature, prior to three further washes and a 1 h incubation with a 1:4000 dilution of anti-human IgG HRP (Invitrogen) at room temperature in the dark. Plates were then washed three times, and 100 µL of SeraCare KPL TMB SureBlue Start solution was added to each well; plate development was halted by the addition of 100 µL SeraCare KPL TMB Stop solution per well. The absorbance at 450nm, with a reference at 650nm, was recorded using a VersaMax microplate reader. For each sample, ELISA endpoint titer was calculated in Graphpad Prism software, using a four-parameter logistic curve fit to calculate the reciprocal serum dilution that yields a corrected absorbance value (450nm-650nm) of 0.2. Log10 endpoint titers are reported.

#### Intracellular cytokine staining (ICS) assay

10^6^ PBMCs/well were re-suspended in 100 µL of R10 media supplemented with CD49d monoclonal antibody (1 µg/mL) and CD28 monoclonal antibody (1 µg/mL). Each sample was assessed with mock (100 µL of R10 plus 0.5% DMSO; background control), pooled S peptides from WA1/2020, B.1.351, B.1.1.7, P.1, and CAL.20C (21^st^ Century Biochemicals) (2 µg/mL), or 10 pg/mL phorbol myristate acetate (PMA) and 1 µg/mL ionomycin (Sigma-Aldrich) (100µL; positive control) and incubated at 37°C for 1 h. After incubation, 0.25 µL of GolgiStop and 0.25 µL of GolgiPlug in 50 µL of R10 was added to each well and incubated at 37°C for 8 h and then held at 4°C overnight. The next day, the cells were washed twice with DPBS, stained with aqua live/dead dye for 10 mins and then stained with predetermined titers of mAbs against CD279 (clone EH12.1, BB700), CD4 (clone L200, BV711), CD27 (clone M-T271, BUV563), CD8 (clone SK1, BUV805), CD45RA (clone 5H9, APC H7) for 30 min. Cells were then washed twice with 2% FBS/DPBS buffer and incubated for 15 min with 200µL of BD CytoFix/CytoPerm Fixation/Permeabilization solution. Cells were washed twice with 1X Perm Wash buffer (BD Perm/Wash™ Buffer 10X in the CytoFix/CytoPerm Fixation/ Permeabilization kit diluted with MilliQ water and passed through 0.22µm filter) and stained with intracellularly with mAbs against Ki67 (clone B56, BB515), IL21 (clone 3A3-N2.1, PE), CD69 (clone TP1.55.3, ECD), IL10 (clone JES3-9D7, PE CY7), IL13 (clone JES10-5A2, BV421), IL4 (clone MP4-25D2, BV605), TNF-α (clone Mab11, BV650),, IL17 (clone N49-653, BV750), IFN-γ (clone B27; BUV395), IL2 (clone MQ1-17H12, BUV737), IL6 (clone MQ2-13A5, APC), CD3 (clone SP34.2, Alexa 700), for 30 min. Cells were washed twice with 1X Perm Wash buffer and fixed with 250µL of freshly prepared 1.5% formaldehyde. Fixed cells were transferred to 96-well round bottom plate and analyzed by BD FACSymphonyTM system.

### SUPPLEMENTARY TABLE

**Supplementary Table S1.** Median pseudovirus neutralizing antibody titers to SARS-CoV-2 variants.

|  | <u>**DAY 29\***</u> | <u>**DAY 239\*\***</u> |
| --- | --- | --- |
| WA1/2020 | 272 | 184 |
| D614G | 167 | 158 |
| B.1.1.7 (ALPHA) | 60 | 147 |
| B.1.617.1 (KAPPA) | 49 | 171 |
| B.1.617.2 (DELTA) | 39 | 107 |
| P.1 (GAMMA) | 28 | 129 |
| B.1.429 (EPISLON) | 27 | 87 |
| B.1.351 (BETA) | <20 | 62 |

**\*Based on 20 participants who received one immunization with Ad26.COV2.S (Fig. 1B).**

### SUPPLEMENTARY FIGURE LEGEND

**Supplementary Figure S1.**
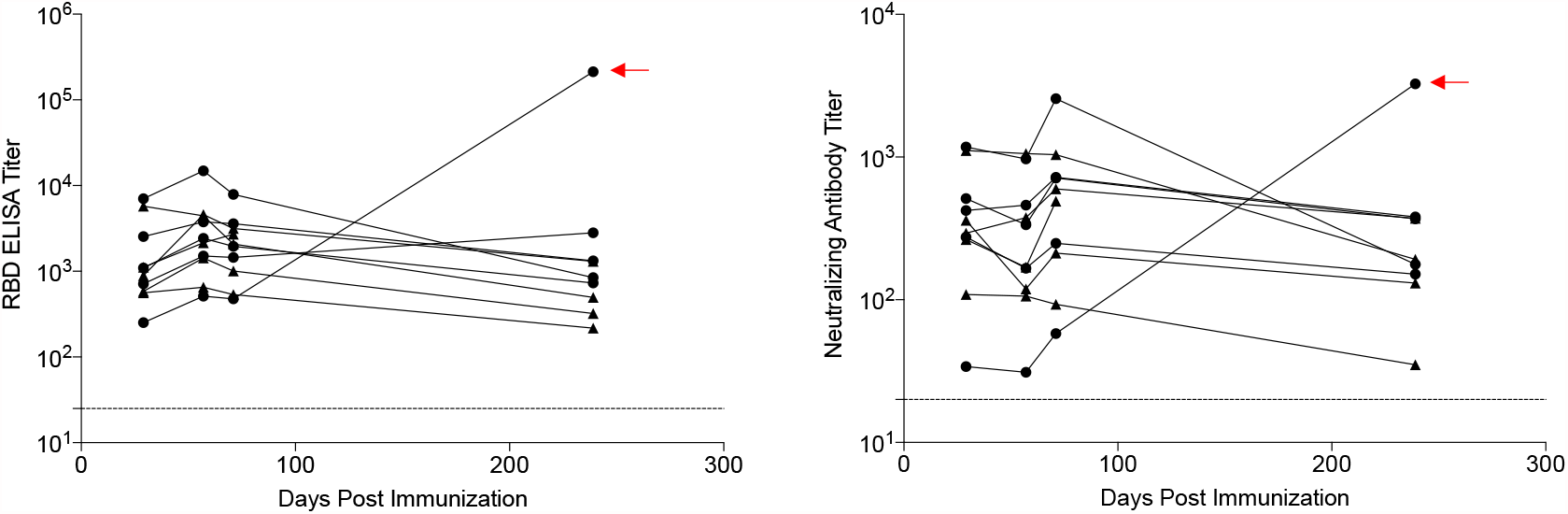
Durability of humoral immune responses following single-shot Ad26.COV2.S vaccination. SARS-CoV-2 WA1/2020 receptor binding domain (RBD)-specific binding antibodies by ELISA and pseudovirus neutralizing antibody assays on days 29, 57, 71 or 85, and 239 restricted to individuals who received single-shot Ad26.COV2.S vaccination. Dotted lines reflect lower limits of quantitation. Filled circles, 10^11^ vp (single dose); filled triangles, 5×10^10^ vp (single dose). Red arrows highlight one individual who developed breakthrough SARS-CoV-2 infection between days 71 and 239.

